# Mental symptoms in Post-COVID Syndrome (PCS) and Post-COVID-19 vaccination syndrome (PCVS): results of a representative population survey

**DOI:** 10.1101/2025.04.02.25325121

**Authors:** Andreas Czaplicki, Leonie Hipper, Ulrich Hegerl

**Affiliations:** German Foundation for Depression and Suicide Prevention, Leipzig, Germany; Department of Psychiatry, Psychosomatic Medicine and Psychotherapy, University Hospital Frankfurt, Goethe University Frankfurt (Goethe Research Professorship), Frankfurt am Main, Germany

## Abstract

Similarities between mental symptoms in Post-COVID syndrome (PCS) and those in Post-COVID-19 vaccination syndrome (PCVS) have been observed. Based on a representative survey of the population aged 18 to 69 years in Germany (n = 4,628), the epidemiologies of both PCS and PCVS were studied. Mental symptoms of PCS (fatigue/exhaustion/weakness, reduced performance, concentration and memory problems, sleep disorders and depressed mood) were reported by 12.1% and mental symptoms of PCVS were reported by 12.6% of all respondents. Concerning PCS, mental symptoms were reported more often by females compared to males (13.5% versus 10.7%), whereas no gender differences were found for PCVS. With the increasing number of booster vaccinations, mental PCVS decreased from 20.8% with one vaccination to 8.9% with four or more vaccinations. Overall, non-mRNA vaccines did not lead significantly to more mental symptoms than mRNA vaccines (12.9% versus 12.5%) but did differ concerning mental symptom patterns: concentration and memory problems (8.2% versus 3.3%), impaired performance (5.7% versus 4.2%) and sleep disorders (6.6% versus 3.2%) were reported more frequently in non-mRNA vaccines. The reported rate of mental PCVS being 12.6% is clearly higher than the 0.5% recorded by official documentation systems (Paul-Ehrlich-Institut (PEI)), suggesting a major underreporting. The data indicate that mental symptoms occurring in both PCS and PCVS are common, making the differential diagnosis between PCS and PCVS, as well as between both PCS and PCVS and independent mental disorders such as depressive disorders, an important but challenging task.

## Introduction

Mental symptoms such as fatigue and cognitive impairment are common in both Post-COVID syndrome (PCS) and Post-COVID-19 vaccination syndrome (PCVS) [1].

PCS is defined as otherwise unexplained symptoms that are present three months after COVID-19 infection and lasting for at least another three months [2]. The proportions of people affected by PCS differ in various population-based surveys and range from 7.3% to 28.5% [3 - 6].

In a systematic review, insomnia was the most common symptom in people with PCR-confirmed COVID infection (pooled prevalence = 27.4%), followed by fatigue (24.4%) and anxiety (19.1%) [7]. Other studies also report memory problems [8], cognitive impairment (e.g. ‘brain fog’), anxiety and low mood [9, 10].

PCVS comprises harmful and unintended reactions to the vaccinations. In the general population, the term ‘post-vac’ is often used for long-lasting side effects following COVID-19 vaccination, whereby the term ‘post-vac’ is not a medically defined term for a disease and is not clearly defined [11]. Compared to the large number of vaccinations, the number of documented side effects is low; the EMA (European Medicines Agency) states the proportion of side effects being 0.2% [12], while the Paul Ehrlich Institute (PEI) reported 0.52% suspected cases of adverse reactions or vaccination complications following COVID-19 vaccines [11].

While there are numerous studies on the somatic side effects of COVID-19 vaccinations [13 - 17], studies focussing on the mental side effects of COVID-19 vaccination are rare. Fatigue has been reported as a systemic side effect of post-COVID-19 vaccination [18, 11]. In addition to the case studies [19], the majority of studies focus on very specific cohorts (e.g. health care providers) [20 - 22] but, as far as we can see, none are population based.

Whether and how mental PCVS is related to the number of vaccinations a person has received has not yet been investigated, as far as we know.

To our knowledge, there are no comprehensive data on the mental symptoms of PCVS following vaccination with mRNA vaccines or with non-mRNA vaccines. In Germany, fatigue has been found to occur slightly more frequently with non-mRNA vaccines than with mRNA vaccines. Although only 8.5% of all COVID vaccinations in Germany were given using non-mRNA vaccines, 10.6% of all cases of chronic fatigue syndrome occurred following these vaccinations. The remaining 91.5% of all COVID vaccinations administered were with mRNA vaccines, however, only 89.4% of all cases of chronic fatigue syndrome occurred following these vaccinations [11, 23].

However, the data on both PCS and PCVS are subject to the risk of bias due to the way in which they are recorded. Spontaneous reports from the public are likely to be biased by underreporting although, opposing this, increased public attention may also lead to an overreporting. Furthermore, the effort required by physicians to report an adverse reaction is likely to lead to significant underreporting. Having these limitations in mind, population-based representative surveys on both PCS and PCVS provide additional information concerning the epidemiology and symptomatology of these syndromes.

In this paper, results of a representative survey on the epidemiologies and symptomatologies of both PCS and PCVS in the adult German population were presented. The following specific research questions were addressed:

1. How common are mental symptoms in people claiming to suffer from PCS and are there subgroups with an enhanced risk?
2. How common are mental symptoms in people claiming to suffer from PCVS and are there subgroups with an enhanced risk?
3. Does the number of vaccinations have an effect on the PCVS risk?
4. Are there differences concerning PCVS between mRNA and non-mRNA vaccines?

## Materials and methods

### Study population and sample characteristics

A representative online survey of the German resident population aged 18 to 69 years (n = 4,628) was conducted from 28^th^ August to 8^th^ September 2023. A multiple stratified quota sample using the interleaved characteristics of gender, age and state/province groups was assessed. The basis for the specifications in the quota cells were the current population updates of the Federal Statistical Office. The study protocol has been reviewed by the Ethics Committee of the Department of Medicine at Goethe University Frankfurt (file number 2023-1174). In a certificate dated 17^th^ February 2023, it was confirmed that there was no obligation to obtain ethics approval for the anonymous data collection.

### Measures

#### PCS

Respondents were asked whether, in their own opinion, they had suffered from PCS using the following wording: "In your opinion, have you suffered from long COVID? Long COVID refers to symptoms that persist, worsen or reappear more than four weeks after infection with the coronavirus, for which there is no other explanation. Long-COVID also includes post-COVID syndrome, which refers to symptoms that appear or persist three months or longer after a coronavirus infection." The possible answers were ‘Yes’, ‘No’ or ‘Don’t know’. The following complaints were considered to be mental symptoms: restricted performance, tiredness/fatigue/weakness, concentration and memory problems, sleep disorders or a depressed mood/depression.

#### PCVS

Respondents who were vaccinated against COVID were asked about PCVS with the following question: "In your opinion, have you experienced any vaccination side effects (lasting consequences) from a COVID vaccination?" The possible answers here were also ‘Yes’, ‘No’ or ‘Don’t know’.

The symptoms of PCVS were queried as follows: “What symptoms do you suffer from? (multiple answers possible): shortness of breath, tiredness/fatigue/weakness, concentration and memory problems, reduced performance, muscle pain, cough/coughing irritation, sleep disorders, palpitations/heart palpitations, loss/change of taste and/or smell, headache, chest pain, tightness in the chest, joint pain, depressed mood/depression, other complaints.” These symptoms were described as symptoms of Long COVID by >= 50% of respondents in a Delphi study. Participants in the Delphi study included clinical researchers, patients, members of the WHO Research Working Group for the Clinical Characterization and Management of COVID-19, members of the WHO Clinical Network for COVID-19, members of a Long COVID SOS patient group, and clinicians and patients nominated by WHO officials [24].

#### Number of vaccinations

The number of COVID-19 vaccinations received was queried as follows: “Have you been vaccinated against COVID?” The possible answers were: ‘yes, once’, ‘twice’, ‘three times’, ‘four times’, ‘five times’, ‘more than five times’, ‘no’ and ‘don’t know’.

#### Type of vaccine (mRNA and non-mRNA)

The following question was used to determine which vaccine was used for the vaccinations: “Which vaccine(s) were you vaccinated with? (multiple answers possible).” Possible answers were the ‘Comirnaty from BioNTech/Pfizer (mRNA vaccine)’, ‘Spikevax from Moderna (mRNA vaccine)’, ‘Jcovden from Janssen-Cilag’, ‘Vaxzevria from AstraZeneca’, ‘VidPrevtyn from Sanofi’, ‘Nuvaxovid from Novavax’, ‘COVID-19 Vaccine Valneva from Valneva’, ‘Bimervax from HIPRA’, ‘Sputnik’ and ‘other and namely (open information)’.

### Statistics

We present descriptive results of the surveys. The data were weighted according to age, gender and federal state using data from official statistics on the structure of the German population. Thus, the sample corresponds to the population composition in 18-69 years age group analysed with regard to the three characteristics mentioned above. The occurrence of PCS was analysed for the subgroups of gender and age group. The occurrence of PCVS was analysed for the subgroups of gender, age group, vaccine and number of vaccinations. Odds ratios (OR) with a reference category, and 95% confidence intervals were calculated for group comparisons. All analyses were carried out using SPSS version 29.0.2.0.

## Results

### Epidemiology of mental symptoms of PCS

In the survey, 12.1% (n = 560) of all respondents mentioned at least one of the mental symptoms of PCS that we presented in the survey. The prevalence of different symptoms are shown in Figure 1.

**Fig 1.**
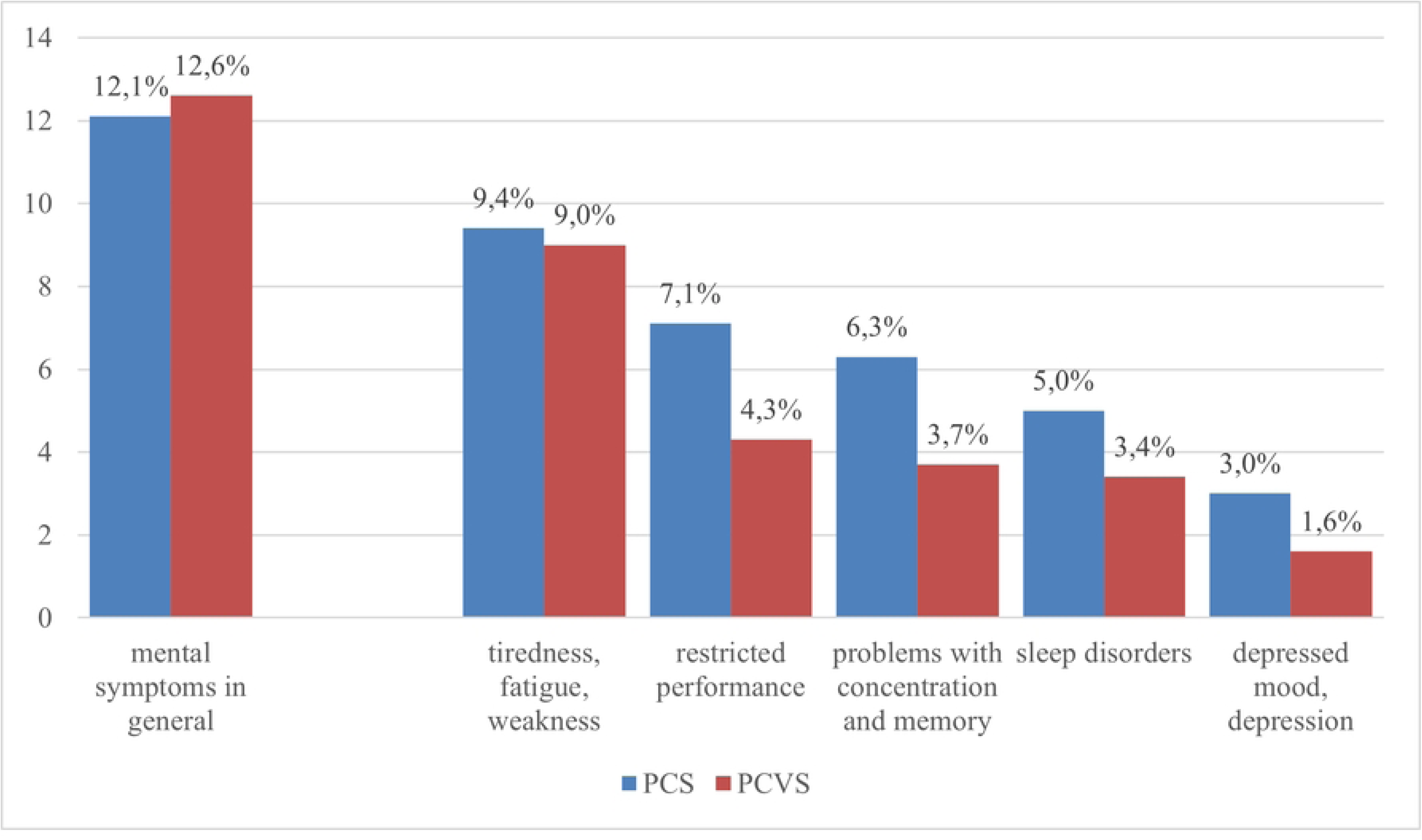
Reported mental symptoms in both the context of PCS and PCVS. (summary and for 5 symptoms separately). Blue bar: all respondents (N = 4,628); red bar: all persons with vaccination (N = 4,049)

Females reported mental symptoms of PCS more often (13.5%; n = 311) compared to males (10.7%; n = 249, p = 0.0032). People aged 18 to 29 years were the most likely to report psychological symptoms of PCS (14.0%, n = 128), while people aged 60 to 69 years were the least likely to report psychological symptoms of PCS (9.4%, n = 87). In relation to the gender of the respondents, this result is also evident in the five different symptoms that the respondents were asked to choose from. All symptoms were reported significantly more frequently by women than by men. Sleep disorders considered to be due to PCS were significant but also slightly higher in the 50-59 years age group (6.2%, p = 0.0269; S1 and S2 Tables).

**S1 Table.** Reported mental problems of PCS by gender and age (self-report, n=4,628, general population 18-69 years).

|  |  | *n/total* | %w | [95% CI] |  | OR | [95% CI] |  | *p* |
| --- | --- | --- | --- | --- | --- | --- | --- | --- | --- |
| gender | total | 560/4,628 | 12.1 | 11.1 | 13.2 |  |  |  |  |
|  | *female (=Ref.)* | 311/2,299 | 13.5 | 12.1 | 15.1 |  |  |  |  |
|  | **male** | **249/2,329** | **10.7** | **9.4** | **12.1** | **0.77** | **0.64** | **0.91** | **0.0032** |
| age | 18-29 y | 128/913 | 14.0 | 11.7 | 16.7 | 1.15 | 0.87 | 1.51 | 0.3217 |
|  | *30-39 y (=Ref.)* | 113/908 | 12.4 | 10.3 | 15.0 | 1.00 |  |  |  |
|  | 40-49 y | 95/835 | 11.4 | 9.2 | 13.9 | 0.90 | 0.68 | 1.21 | 0.4923 |
|  | 50-59 y | 120/1,048 | 11.5 | 9.5 | 13.7 | 0.91 | 0.69 | 1.20 | 0.4984 |
|  | **60-69 y** | **87/924** | **9.4** | **7.5** | **11.6** | **0.73** | **0.54** | **0.98** | **0.0382** |

Percentages weighted by age, gender and federal state in Germany (%w, [95% confidence interval]) and odds ratios (OR). From N=4,628 people (general population 18-69 years). Significant differences are highlighted in bold. The reference categories are marked with (=Ref.)

**S2 Table.**
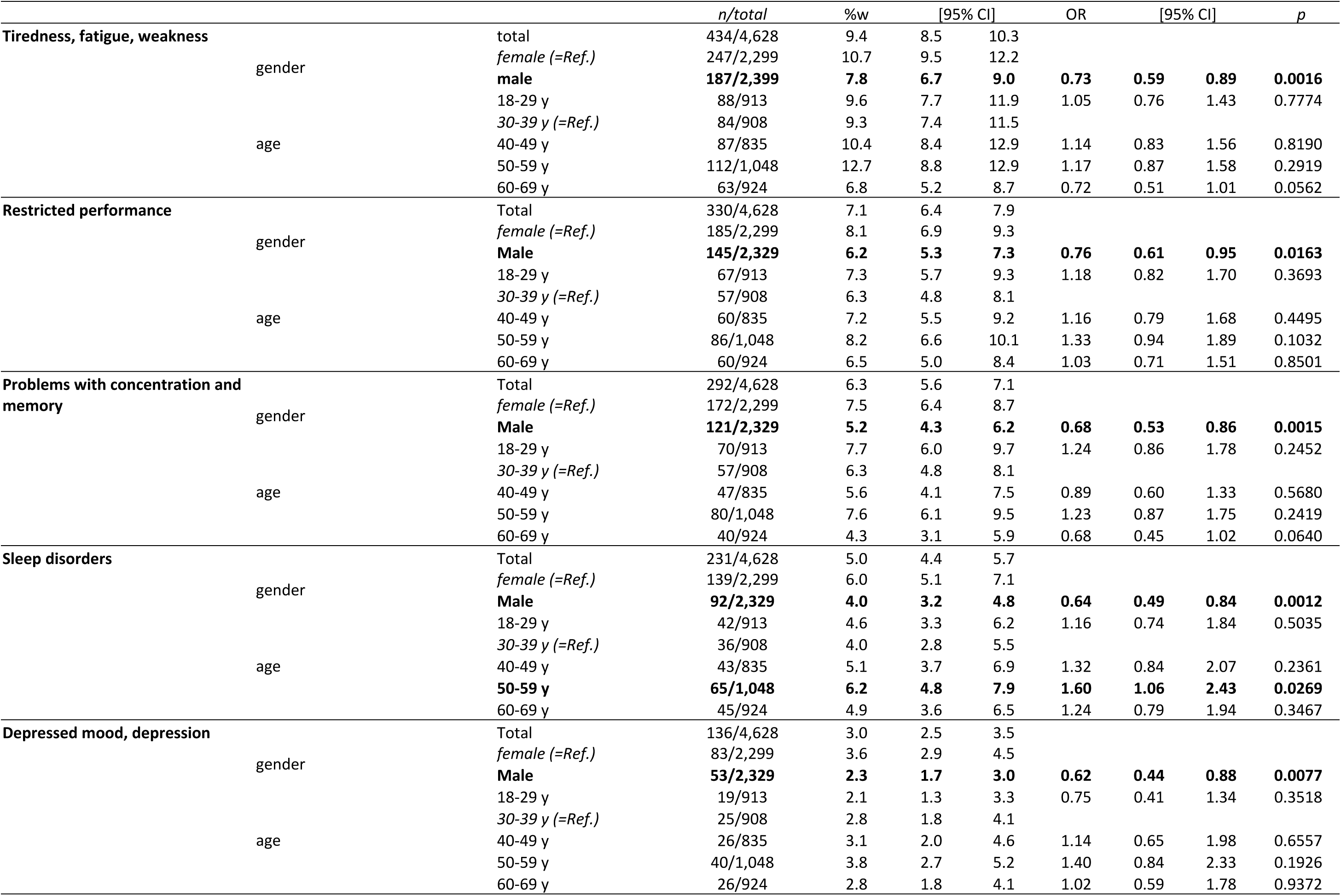
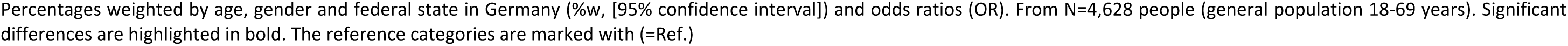
Reported mental symptoms of PCS (differentiated) by gender and age (self-report, n=4,628, general population 18-69 years).

### Epidemiology of mental vaccination side effects

In the survey, 88.1% (n = 4,079; 87.9% males, 88.4% females) of respondents stated that they had been vaccinated against SARS-CoV2; in the 18-29 years it was 87.8% (n = 802), in the age group of 30-39 years it was 87.2% (n = 792), for the group aged 40-49 years it was 85.2% (n = 711), while in the age group 50-59 years it was 88.6% (n = 929), and in the group aged 60-69 years it was 91.5% (n = 845).

Of the respondents, 12.6% (n = 512) of respondents reported mental side effects of the COVID-19 vaccination. The following symptoms were again summarised here: restricted performance, tiredness/fatigue/weakness, concentration and memory problems, sleep disorders or a depressed mood/depression.

Mental problems as side effects of COVID vaccination were mentioned for the individual symptoms: tiredness/fatigue/weakness (9.0%), restricted performance (4.3%), concentration and memory problems (3.7%), sleep disorders (3.4%) or a depressed mood/depression (1.5%). This order of symptoms was the same as for PCS (Fig 1).

Regarding mental PCVS, no gender differences were found (S3 Table). Only the tiredness/fatigue/weakness symptoms were reported less frequently by men (7.8%) than by women (10.2%, p = 0.0069). Age-related differences were only found for the problems with concentration and memory; here, older groups reported corresponding side effects less frequently (S4-S8 Tables).

**S3 Table.**
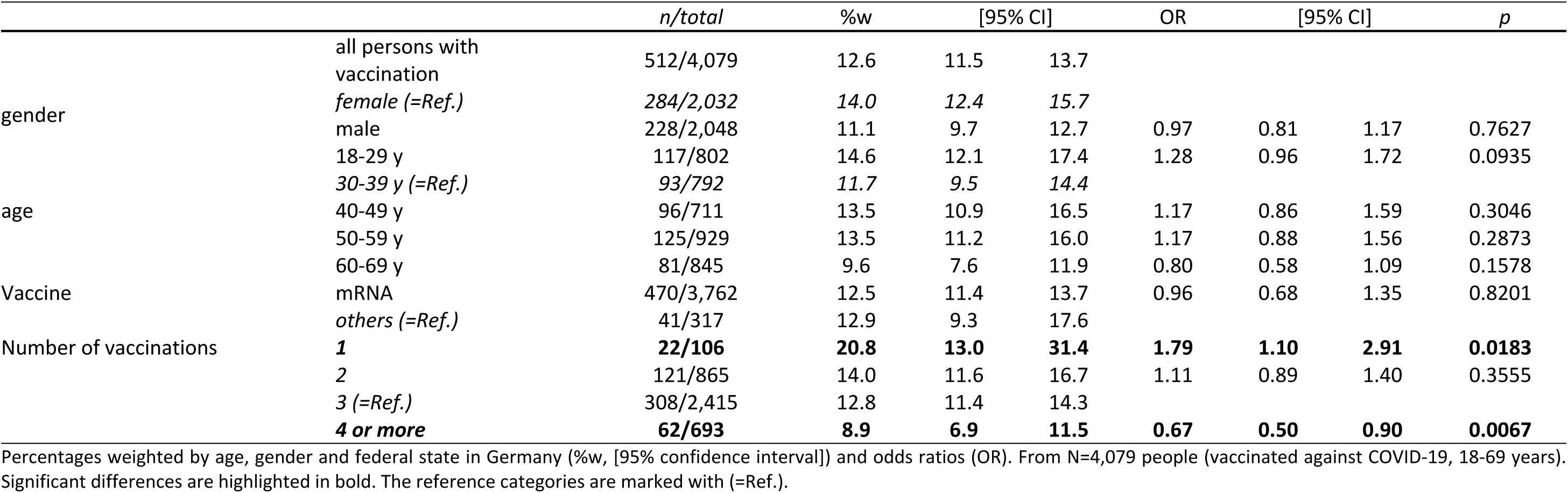
Reported mental side effects of COVID-19 vaccination (self-report, n=4.079 respondents with vaccination, population 18-69 years) by gender and age.

**S4 Table.**
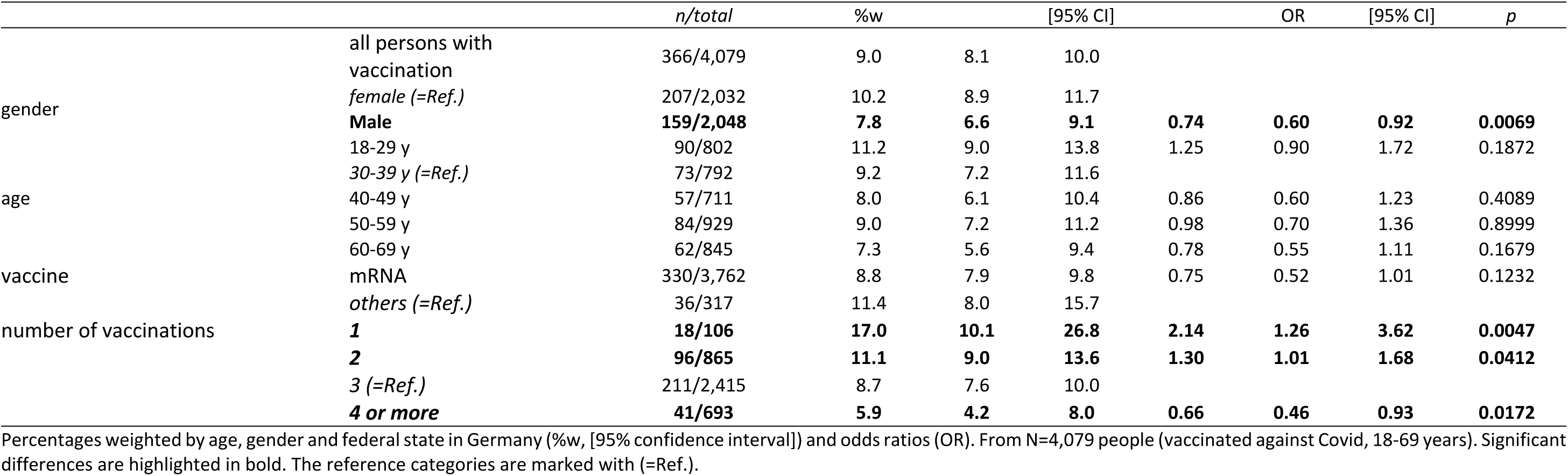
Reported tiredness, fatigue, weakness as side effects of covid vaccination by gender, age, vaccine and number of vaccinations (self-report, n=4.079 respondents with vaccination, population 18-69 years).

**S5 Table.**
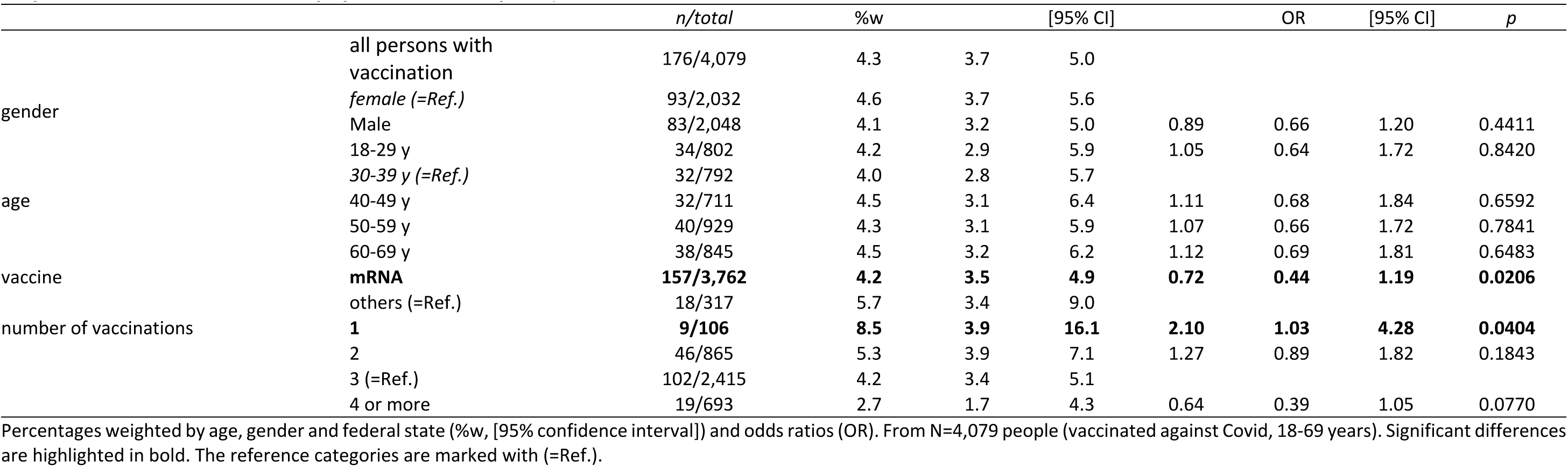
Restricted performance as side effects of covid vaccination by gender, age, vaccine and number of vaccinations (self-report, n=4.079 respondents with vaccination, population 18-69 years).

**S6 Table.**
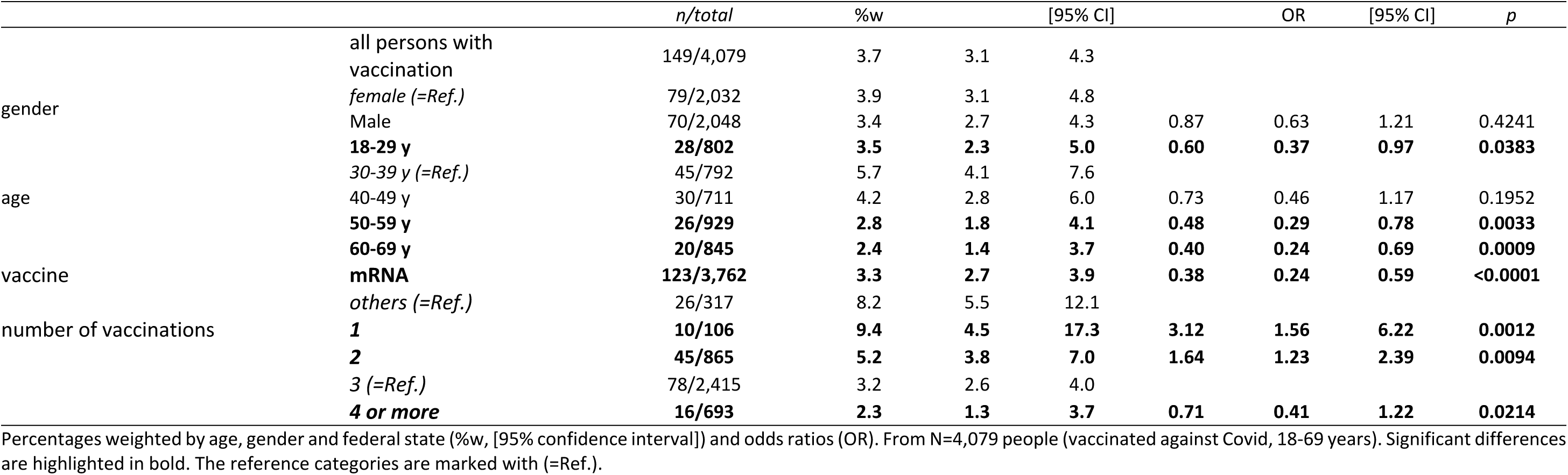
Problems with concentration and memory as side effects of covid vaccination by gender, age, vaccine and number of vaccinations (self-report, n=4.079 respondents with vaccination, population 18-69 years).

**S7 Table.**
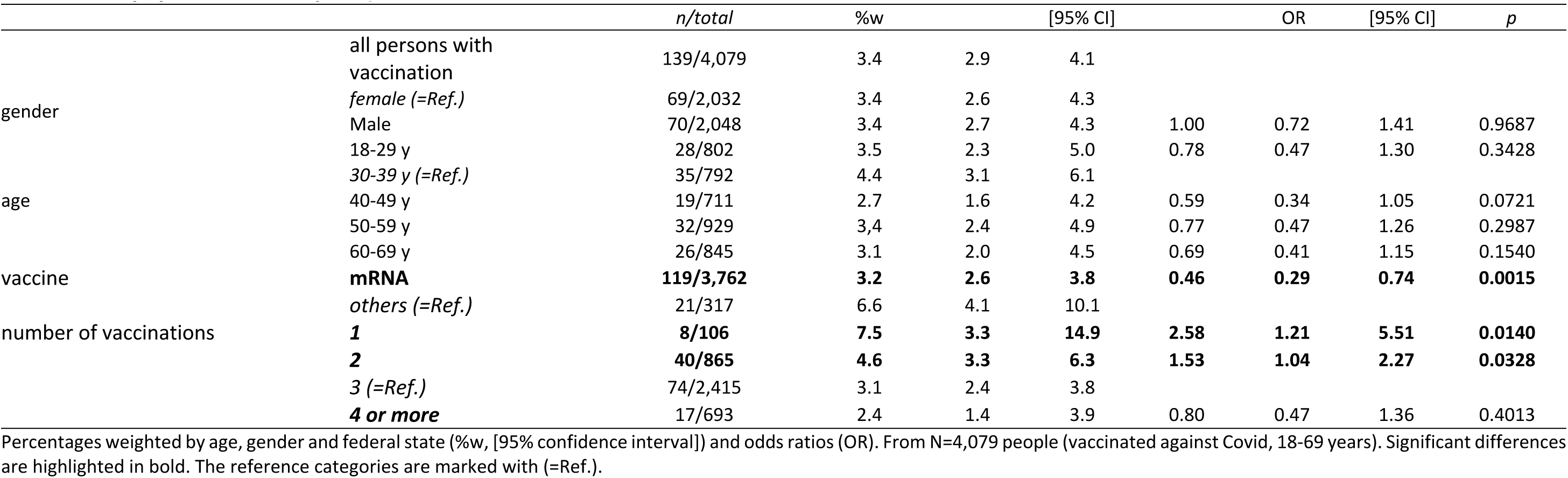
Sleep disorders as side effects of covid vaccination by gender, age, vaccine and number of vaccinations (self-report, n=4.079 respondents with vaccination, population 18-69 years).

**S8 Table.**
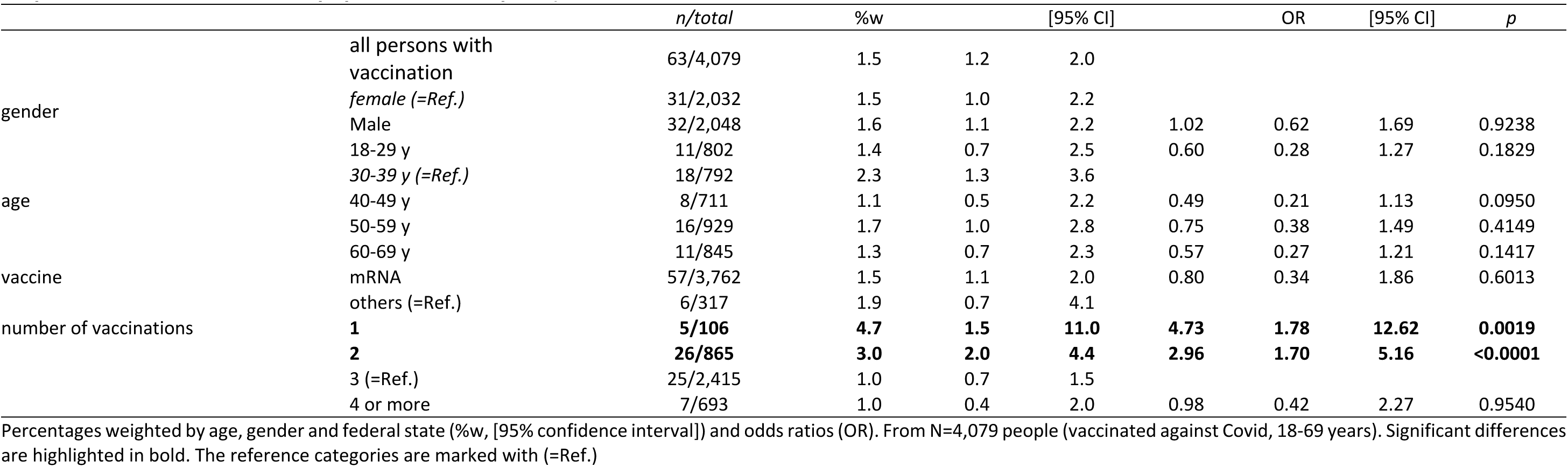
Depressed mood/ depression as side effects of covid vaccination by gender, age, vaccine and number of vaccinations (self-report, n=4.079 respondents with vaccination, population 18-69 years).

### Effects of the number of vaccinations on mental symptoms

Of those vaccinated, 2.6% (n = 106) reported one vaccination, 21.2% (n = 865) reported two vaccinations, 59.2% (n = 2,415) reported three vaccinations, and 17.0% (n = 693) reported four or more vaccinations. The reported mental side effects of the COVID vaccination decreased with the number of vaccinations: 20.8% (n = 22) with one vaccination, decreasing to 8.9% (n = 62) with four or more vaccinations.

The number of COVID vaccinations received showed significant effects for almost the mental symptoms surveyed. Respondents with either one or two vaccinations reported these symptoms more frequently compared to respondents having had four or more vaccinations (S3-S7 Tables).

### Differences in reported side effects between mRNA and non-mRNA vaccines

Overall, 92.2% (n = 3,762) of all vaccinated persons were vaccinated at least once with an mRNA vaccine. Of those vaccinated, 81.6% received a vaccination with the "Comirnaty" vaccine from BioNTech/Pfizer, 32.1% reported at least one vaccination with "Spikevax" from Moderna, 17.7% with "Vaxzevria" from AstraZeneca, and 4.9% with the vaccine "Jcovden" from Janssen-Cilag. All other vaccines were administered only rarely. A few respondents also reported vaccinations with unauthorised vaccines or vaccines with other indications, e.g. Bionical, Asenica, Sinovax.

Types of vaccine (mRNA or non-mRNA) showed no significant differences when all mental symptoms surveyed were taken together. However, differences could be seen when differentiating between the various mental symptoms. Recipients of non-mRNA vaccines reported restricted performance more frequently (5.7%) than recipients of mRNA vaccines (4.2%, p = 0.0206); the same applied to problems with concentration and memory (8.2% cf. 3.3%, p < 0.0001) and sleep disorders (6.6% cf. 3.2%, p = 0.0015).

## Discussion

In our study, 12.1% of the general population reported that they had experienced mental symptoms of PCS. This value is lower than in other studies, however, in those studies, the values were determined in a clinical setting and, therefore, represented a specific population, whereas our values relate to the general population. Here, as there, fatigue proved to be the most prevalent mental symptom of PCS. When assessing gender differences in PCS, we found that women were more likely to report mental symptoms of PCS than men. This finding is in line with studies indicating that women are more frequently affected by PCS than men [25, 26]. In terms of age, younger people were more likely to be affected by PCS than older people.

Mental side effects considered to be PCVS were reported by 12.6% of all of the respondents. No gender-specific differences were found in PCVS. Older respondents were slightly less likely to report problems with concentration and memory.

Our results are significantly higher than the official figures from the Paul-Ehrlich-Institute (0.52%). There are several factors that could, at least, partially explain this difference. Not everyone would have been familiar with the reporting portals and the procedure for reporting side effects. The reporting procedure is also time-consuming which could lead to underreporting by physicians and also by those affected by PCVS and their relatives in the general population. Overall, our figures suggest that more people are affected by PCVS than stated in the official statistics.

An interesting finding is that the pattern of mental symptoms in PCS is at least partially different from that in PCVS. While various symptoms occur more frequently in PCS, fatigue is by far the most relevant symptom in PCVS.

The mRNA and non-mRNA vaccines, in general, did not differ in terms of the rate of mental PCVS. Amongst respondents who were vaccinated with an mRNA vaccine, 12.5% reported at least one of the mental symptoms of PCVS that were available as response options, whilst amongst respondents who were vaccinated with a non-mRNA vaccine, the figure was 12.9%. Respondents reported that some mental symptoms were more common following vaccination with non-mRNA vaccines than after vaccination with mRNA vaccines (concentration/memory problems 8.2% versus 3.3%; impaired performance 5.7% versus 4.2%, and sleep problems 6.6% versus 3.2%). We are not aware of any comparative data in this regard. There were no significant differences in the symptoms of tiredness/fatigue/weakness and depressed mood. With regard to fatigue, this contradicts data from the PEI, in which fatigue occurs slightly more frequently after vaccination with a non-mRNA vaccine than with an mRNA vaccine [27].

People who had received more than two COVID vaccinations reported mental PCVS less often than those receiving two or less. Respondents with four or more vaccinations reported significantly fewer side effects. A likely explanation for this finding is that people experiencing side effects after a first COVID vaccination avoided further vaccinations.

The fact that in the contexts of both PCS and PCVS, mental symptoms are prevalent, as found in this survey, it is likely that this will create problems for clinicians to separate PCS from PCVS. Fatigue and tiredness may result from both reactions to a COVID-19 infection and, for example, immunological changes in the immune system, or a diagnosed or undiagnosed myocarditis due to the COVID-19 vaccination.

The reported mental symptoms that are supposedly part of PCS or PCVS are unspecific and often found in the context of mental disorders, especially depression. This raises the question of how often mental symptoms related to PCS or PCVS are simply symptoms of a hidden and undiagnosed depressive disorder and are, thus unrelated to PCS or PCVS. People suffering from a depressive episode often misattribute their symptoms to causes other than depression and both PCS and PCVS could be explanations at hand. This would be in line with the finding that the mental problems supposedly related to PCS or PCVS are more often reported by women, who also have a higher prevalence of major depression [28, 29].

The data suggest that PCS with mental symptoms as well as mental symptoms in PCVS are common, making the differential diagnosis between PCS and PCVS, as well as between both PCS and PCVS on the one hand, and depressive disorders on the other, difficult.

## Limitations

An obvious limitation is that the data represent the views and the self-assessments of the general population without any health professionals providing a validation and classification concerning the severity of the symptoms. Furthermore information about the duration of the symptoms were lacking in the self-assessments. Personal opinions, convictions and attitudes concerning COVID-19 and the measures against it may have additionally influenced the results. Furthermore, the survey covers only the age group from 18 to 69 years. Over 69-year-olds, a high-risk group for severe COVID cases, were not included in the survey. Moreover, there was a lack of precise information about the time course of the mental symptoms in both PCS and PCVS. The time course of the mental symptoms may be able to help differentiate between PCS and PCVS. While long-COVID is a syndrome that, by definition, persists for three months after an infection, the vaccination side effects usually occur acutely shortly after vaccination. However, there are also vaccination side effects that appear later and, therefore, need to be distinguished from long-COVID [30 - 32].

## Data Availability

The data underlying the results presented in the study are available via the cloud of the German Foundation for Depression and Suicide Prevention via the following link: https://cloud.deutsche-depressionshilfe.de/public/download-shares/iq7vo4PyvIYEpneVmyJdqm5Ox3jyGgCB

## References

1. Bundesärztekammer. Bekanntmachungen: Post-COVID-Syndrom (PCS). Deutsches Ärzteblatt Online. 2022. Available from: https://www.bundesaerztekammer.de/fileadmin/user_upload/BAEK/Themen/Medizin_und_Ethik/BAEK_Stellungnahme_Post-COVID-Syndrom_ONLINE.pdf

2. World Health Organization: A clinical case definition of post COVID-19 condition by a Delphi consensus. Who.int. 2021 Oct 6 [cited 18 March 2024]. World Health Organization [Internet]. Available from: https://iris.who.int/bitstream/handle/10665/345824/WHO-2019-nCoV-Post-COVID-19-condition-Clinical-case-definition-2021.1-eng.pdf?sequence=1

3. Robertson MM, Qasmieh SA, Kulkarni SG, Teasdale CA, Jones HE, McNairy M, et al. The epidemiology of long coronavirus disease in US adults. Clin Infect Dis. 2023 May 3;76(9):1636–45. doi:10.1093/cid/ciac961

4. Ballering AV, van Zon SKR, Olde Hartman TC, Rosmalen JGM. Persistence of somatic symptoms after COVID-19 in the Netherlands: an observational cohort study. Lancet. 2022 Aug 6;400(10350):452–461. doi: 10.1016/S0140-6736(22)01214-4

5. Mogensen I, Ekström S, Hallberg J, Georgelis A, Melén E, Bergström A, et al. Post covid-19 symptoms are common, also among young adults in the general population. Sci Rep. 2023 Jul 12;13(1). doi:10.1038/s41598-023-38315-2

6. Peter RS, Nieters A, Kräusslich H-G, Brockmann SO, Göpel S, Kindle G, et al. Post-acute sequelae of covid-19 six to 12 months after infection: Population Based Study. BMJ. 2022 Oct 13. doi:10.1136/bmj-2022-071050

7. Badenoch JB, Rengasamy ER, Watson C, Jansen K, Chakraborty S, Sundaram RD, et al. Persistent neuropsychiatric symptoms after COVID-19: A systematic review and meta-analysis. Brain Commun. 2021 Dec 17;4(1). doi:10.1093/braincomms/fcab297

8. Chen C, Haupert SR, Zimmermann L, Shi X, Fritsche LG, Mukherjee B. Global prevalence of post-coronavirus disease 2019 (COVID-19) condition or long covid: A meta-analysis and systematic review. J Infect Dis. 2022 Apr 16;226(9):1593–607. doi:10.1093/infdis/jiac136

9. Lopez-Leon S, Wegman-Ostrosky T, Perelman C, Sepulveda R, Rebolledo PA, Cuapio A, et al. More than 50 long-term effects of COVID-19: A systematic review and meta-analysis. Sci Rep. 2021 Aug 9;11(1). doi:10.1038/s41598-021-95565-8

10. Raveendran AV, Jayadevan R, Sashidharan S. Long COVID: An overview. Diabetes Metab Syndr. 2021 Apr 20;15(3):869–75. doi:10.1016/j.dsx.2021.04.007

11. Mentzer D, Oberle B, Streit R, Weisser K, Keller-Stanislawski B. Sicherheitsprofil der COVID-19-Impfstoffe – Sachstand. Bulletin zur Arzneimittelsicherheit. 2023 Mar 31;14:12–29

12. European Medicines Agency. Safety of COVID-19 vaccines. 2023 [cited 10 Feb 2025]. European Medicines Agency [Internet]. Available from: https://www.ema.europa.eu/en/human-regulatory-overview/public-health-threats/coronavirus-disease-covid-19/covid-19-medicines/safety-covid-19-vaccines

13. Kouhpayeh H, Ansari H. Adverse events following covid-19 vaccination: A systematic review and meta-analysis. Int Immunopharmacol. 2022 May 30;109:108906. doi:10.1016/j.intimp.2022.108906

14. Meo SA, Bukhari IA, Akram J, Meo AS, Klonoff DC. COVID-19 vaccines: comparison of biological, pharmacological characteristics and adverse effects of Pfizer/BioNTech and Moderna Vaccines. Eur Rev Med Pharmacol Sci. 2021 Feb 1;25(3):1663–9. doi:10.26355/eurrev_202102_24877

15. Rabail R, Ahmed W, Ilyas M, Rajoka MS, Hassoun A, Khalid AR, et al. The side effects and adverse clinical cases reported after COVID-19 immunization. Vaccines. 2022 Mar 22;10(4):488. doi:10.3390/vaccines10040488

16. Medeiros KS, Costa AP, Sarmento AC, Freitas CL, Gonçalves AK. Side effects of covid-19 vaccines: A systematic review and meta-analysis protocol of randomised trials. BMJ Open. 2022 Feb 24;12(2). doi:10.1136/bmjopen-2021-050278

17. Beatty AL, Peyser ND, Butcher XE, Cocohoba JM, Lin F, Olgin JE, et al. Analysis of COVID-19 vaccine type and adverse effects following vaccination. JAMA Netw Open. 2021 Dec 22;4(12). doi:10.1001/jamanetworkopen.2021.40364

18. Loosen SH, Bohlken J, Weber K, Konrad M, Luedde T, Roderburg C, et al. Factors associated with non-severe adverse reactions after vaccination against SARS-COV-2: A cohort study of 908,869 outpatient vaccinations in Germany. Vaccines. 2022 Apr 6;10(4):566. doi:10.3390/vaccines10040566

19. Roberts K, Sidhu N, Russel M, Abbas MJ. Psychiatric pathology potentially induced by covid-19 vaccine. Prog Neurol Psychiatry. 2021 Nov 8;25(4):8–10. doi:10.1002/pnp.723

20. Al-Obaidy LM, Attash HM, Al-Qazaz HK. Depression, anxiety and stress after COVID-19 vaccination: A retrospective cross-sectional study among health care providers. Pharm Pract. 2022 Sept 23;20(3):01–8. doi:10.18549/pharmpract.2022.3.2689

21. Bareiß A, Uzun G, Mikus M, Becker M, Althaus K, Schneiderhan-Marra N, Fürstberger A, Schwab JD, Kestler HA, Holderried M, Martus P, Schenke-Layland K, Bakchoul T: Vaccine Side Effects in Health Care Workers after Vaccination against SARS-CoV-2: Data from TüSeRe:exact Study Viruses 15, no. 1: 65. doi:10.3390/v15010065

22. Rifai A, Wu WC, Tang YW, Lu MY, Chiu PJ, Strong C, et al. Psychological distress and physical adverse events of covid-19 vaccination among healthcare workers in Taiwan. Vaccines. 2023 Jan 5;11(1):129. doi:10.3390/vaccines11010129

23. Robert Koch Institut: Digitales Impfquotenmonitoring zur COVID-19-Impfung (bis 30.6.2024). Tabelle mit den gemeldeten Impfungen nach Bundesländern und Impfquoten nach Altersgruppen. 2024, Robert Koch Institut [Internet]. [cited 20 February 2024]. Available from: https://www.rki.de/DE/Themen/Infektionskrankheiten/Impfen/Impfungen-A-Z/COVID-19/Impfquoten/Impfquotenmonitoring.xlsx?blob=publicationFile&v=3

24. Soriano JB, Murthy S, Marshall JC, Relan P, Diaz JV. A clinical case definition of post-covid-19 condition by a delphi consensus. Lancet Infect Dis. 2021 Dec 21;22(4):e102–7. doi:10.1016/s1473-3099(21)00703-9

25. Chen C, Haupert SR, Zimmermann L, Shi X, Fritsche LG, Mukherjee B. Global prevalence of post-coronavirus disease 2019 (COVID-19) condition or long covid: A meta-analysis and systematic review. J Infect Dis. 2022 Apr 16;226(9):1593–607. doi:10.1093/infdis/jiac136

26. Thompson EJ, Williams DM, Walker AJ, Mitchell RE, Niedzwiedz CL, Yang TC, et al. Long Covid Burden and risk factors in 10 UK longitudinal studies and Electronic Health Records. Nat Commun. 2022 Jun 28;13(1). doi:10.1038/s41467-022-30836-0

27. Paul-Ehrlich-Institut. Sicherheitsbericht des Paul-Ehrlich-Instituts zur COVID-19-Impfkampagne in Deutschland. 2022 [cited 10 Feb 2025]. Paul-Ehrlich-Institut [Internet]. Available from: https://www.pei.de/SharedDocs/Downloads/DE/newsroom/dossiers/sicherheitsberichte/sicherheitsbericht-27-12-20-bis-30-06-22.pdf?blob=publicationFile&v=6

28. Gutiérrez-Rojas L, Porras-Segovia A, Dunne H, Andrade-González N, Cervilla JA. Prevalence and correlates of major depressive disorder: A systematic review. BJP. 2020 Aug 2;42(6):657–72. doi:10.1590/1516-4446-2020-0650

29. Salk RH, Hyde JS, Abramson LY. Gender differences in depression in representative national samples: Meta-analyses of diagnoses and symptoms. Psychological Bulletin. 2017 Apr 17;143(8):783–822. doi:10.1037/bul0000102

30. Scholkmann F, May C-A. Covid-19, post-acute covid-19 syndrome (PACS, “long covid”) and post-covid-19 vaccination syndrome (PCVS, “post-covidvac-syndrome”): Similarities and differences. Pathol Res Pract. 2023 May 3;246:154497. doi:10.1016/j.prp.2023.154497

31. Riad A, Pokorná A, Attia S, Klugarová J, Koščík M, Klugar M. Prevalence of COVID-19 vaccine side effects among healthcare workers in the Czech Republic. J Clin Med. 2021 Apr 1;10(7):1428. doi:10.3390/jcm10071428

32. Klugar M, Riad A, Mekhemar M, Conrad J, Buchbender M, Howaldt H-P, et al. Side effects of mrna-based and viral vector-based COVID-19 vaccines among German healthcare workers. Biology. 2021 Aug 5;10(8):752. doi:10.3390/biology10080752

